# Biomarkers of Parkinson’s Disease: Screening Vital Signs and Routine Blood Tests

**DOI:** 10.1101/2020.05.18.20103085

**Authors:** Hirotaka Iwaki, Hampton L. Leonard, Sara Bandrés-Ciga, Cornelis Blauwendraat, Mary B. Makarious, Sonja W. Scholz, Noriko Nishikawa, Faraz Faghri, Mark Frasier, J. Raphael Gibbs, Andrew B. Singleton, Mike A. Nalls, Dena G. Hernandez

## Abstract

**BACKGROUND:** There is a need for reliable, objective, and easily accessible biomarkers for Parkinson’s disease.

**OBJECTIVES:** The purpose of this study was to screen biomarkers from vital signs and routine blood tests.

**METHODS:** Longitudinal data of up to 7 years of vital signs and routine blood tests from 418 patients with Parkinson’s disease (PD) untreated at baseline and 185 individuals without any neurological disease were analyzed using linear mixed models. We nominated the biomarkers whose main associations with the measurements were significant as differentiating biomarkers. Similarly, we nominated the interaction effects between biomarkers and time from baseline as progression biomarkers. We tested for 49 biomarkers, and multiple comparison was corrected with the false-discovery-rate of 0.05. We further evaluated the potential biomarkers with regard to their importance in diagnosis prediction and their association with sub-scores on the Movement Disorder Society-Unified Parkinson’s Disease Rating Scale (MDS-UPDRS). We also assessed the relationship of the associations using bioinformatics.

**RESULTS:** Heart rate, systolic blood pressure, white blood cell fractions, neutrophil counts, serum albumin, sodium and AST were different between PD and controls. The causality or genetic correlations of these biomarkers to PD were not observed. Chronological changes in height, albumin, hemoglobin, and bicarbonate were different in PD. These biomarkers were associated with MDS-UPDRS sub-scores.

**Conclusions:** In this study, the potential of some easily accessible biomarkers for diagnosis and disease progression was presented. Further investigation of the mechanisms underlying these associations is important for a deeper understanding of the disease and the better management of patients.

## Introduction

There is an ongoing effort to identify reliable and objective biomarkers for Parkinson’s disease (PD) that could assist with diagnosis or change with progression [1]. In addition, studying the mechanisms underlying the associations between biomarkers and a disease is important to understand the disease. One of the most intensively studied candidate biomarkers is the measurement of α-synuclein levels in the cerebrospinal fluid (CSF). The concentration of α-synuclein in the CSF has been consistently reported as being decreased in PD patients [2–4]. It may also be a prognostic marker for motor symptoms [5,6].Because of the large variance, however, the sensitivity (78–88%) and specificity (40–57%) of α-synuclein levels in the CSF are not high enough to use in clinical practice [1]. Additionally, lumbar puncture is not within the range of daily medical procedures. Ideally, biomarkers should be easily accessible and inexpensive.

Here, we screened for biomarkers in vital signs and routine blood laboratory tests, including hematology and chemistry measurements. We were interested in 2 types of biomarkers: (1) biomarkers differentiating cases from controls regardless of time (differentiating biomarkers) and (2) biomarkers associated with the progression of the disease (progression biomarkers). To identify these 2 types of biomarkers, we studied longitudinal data of idiopathic PD patients and controls from the Parkinson’s Progression Marker Initiative (PPMI). Specifically, for differentiating biomarkers, we examined measurements discriminating between PD cases and controls over time. For progression biomarkers, we screened the measurements for which having PD was associated with a different rate of change than not having PD, knowing that PD is a progressive disease. Next, we evaluated the characteristics of the relationships between the nominated blood biomarkers and PD, using genetic approaches. Namely, we conducted linkage disequilibrium (LD) score regressions to assess the genetic correlations between the biomarkers and PD. We also applied 2-sample Mendelian randomization to evaluate causality in the relationships of the associations. We assessed the quality of the nominated differentiating biomarkers and α-synuclein levels in the CSF. We also examined the association between progression biomarkers and sub-scores on the Movement Disorder Society-Unified Parkinson’s Disease Rating Scale (MDS-UPDRS) [7].

## Method

### Study cohort

The participants in this study included 418 patients with PD (cases) and 185 individuals without any neurological disease (controls) from the PPMI. Detailed information regarding the PPMI is available at www.ppmi-info.org. The cases of the study were untreated PD patients who had been diagnosed less than 2 years before enrollment. Most of them had blood samples taken at baseline and then annually. The vital signs were also recorded at each visit. The PPMI is an ongoing study, and we downloaded the data from the website on August 28, 2019 and used the observations from the first 7 years (up to visit 14). The PPMI study was registered at ClinicalTrials.gov (NCT01141023), approved by the institutional review board at each site, and the participants provided written informed consent.

### Laboratory tests

The routine clinical laboratory tests listed below were performed at baseline and annual follow-ups. A central laboratory (Covance Inc., Princeton, NJ, USA) was used to guarantee identical analysis methods and consistent normal ranges and, thus, common interpretation of laboratory changes. The hematology tests included red blood cell counts (TI/L), hemoglobin (g/L), hematocrit (no unit), white blood cell (WBC) counts (GI/L), neutrophil counts (GI/L) and their ratio in WBC (%), eosinophil counts (GI/L) and their ratio in WBC (%), basophil counts (GI/L) and their ratio in WBC (%), monocyte counts (GI/L) and their ratio in WBC (%), lymphocyte counts (GI/L) and their ratio in WBC (%), and platelet counts(GI/L). The chemistry tests included total protein (g/L), albumin (g/L), total bilirubin (μmol/L), alkaline phosphatase (U/L), aspartate aminotransferase (U/L), alanine aminotransferase (U/I), serum sodium (mmol/L), serum potassium (mmol/L), serum bicarbonate (mmol/L), serum chloride (mmol/L), calcium (mmol/L), creatinine (μmol/L), urea nitrogen (mmol/L), serum uric acid (μmol/L), and serum glucose (mmol/L).

The concentration of alpha-synuclein in the CSF samples collected for the PPMI were analyzed using an ELISA assay available commercially from BioLegend (cat # 844101).

### Vital signs and MDS-UPDRS

Heart rate (supine and standing), blood pressure (supine and standing), and oral temperature were measured at every visit. The supine blood pressure and heart rate were determined after 1–3 minutes of quiet rest, and the standing blood pressure and heart rate were determined after 1–3 minutes in the standing position. Weight and height were collected at baseline and annually or bi-annually according to the visit schedule. In addition to the measured vital signs, we created some new outcomes. We examined the differences between the measurements taken in the supine position and those taken in the standing position for systolic blood pressure, diastolic blood pressure, and heart rate. We also examined the ratio of these measurements: We divided the value in the standing position by the value in the supine position. In addition, sub-scores on the MDS-UPDRS part I, part II, and part III at baseline were used in the exploratory analysis of progression markers.

### Statistical analysis

#### Primary analysis: Nominating biomarkers among vital signs and lab measurements

We used linear mixed effect models including terms indexing having or not having PD (PD status), years from baseline, interaction between PD status and years from baseline, sex, age at baseline, BMI, the dosages of PD drugs (levodopa, MAO-B inhibitor, pramipexole, ropinirole, and amantadine), and random terms of intercept and slope for each individual. In this model, the beta coefficient for the PD status term (main term) corresponded to the difference between PD cases and controls over time, while the beta coefficient for the interaction between PD status and years from baseline term(interaction term) corresponded to the difference between PD cases and[8] controls in terms of the rate of change between PD and controls over time. To correct multiple comparison, we calculated qvalues [8] and applied 2-tailed tests at the false discovery rate (FDR) of 0.05 for the number of tested models (49) and nominated differentiating biomarkers whose main term was significant and progression biomarkers whose interaction term was significant. To avoid the impact of outliers on the analyses, we excluded the data points that had outcomes outside of mean +/− 4 SD from the analyses. For the nominated differentiating/progression biomarkers, we also checked the mean difference between PD cases and controls after adjusting for age, sex and BMI; we checked the mean difference between PD cases and controls at 0 (baseline) and at the first-, second-, third-, fourth-, and fifth-year. The model specifications of these analyses are provided in the supplemental materials, and the analyses were conducted in R version 3.6.3.

#### Causal inferences and genetic correlations

We applied LD score regression (LDSC) and 2-sample Mendelian randomization to explore genetic correlations and causal inferences between PD and the nominated biomarkers, respectively. LDSC is based on the premise that single nucleotide polymorphisms (SNPs) in regions of high LD tag a greater proportion of the genome and will therefore show stronger associations, on average, than SNPs in regions of low LD [9]. Using the known LD structure of a reference SNP panel, reliable estimates of shared genetic risk can be estimated. We implemented LDSC v1.0.1, using default settings to calculate overlapping genetic etiologies from the most recent PD genome-wide association study (GWAS) summary statistics [10] and publicly available summary statistics for some of the nominated biomarkers, including neutrophil percentage of white blood cells, neutrophil count, lymphocyte counts, lymphocyte percentage of white blood cells, monocyte percentage of white blood cells, and hemoglobin concentration [11]. Summary statistics for the remaining biomarkers were not available at the time of the analysis.To explore the causal effects of the nominated biomarkers on PD risk, we then implemented 2-sample Mendelian randomization (MR) using MR Base under default settings [12]. We undertook harmonization was to rule out strand mismatches and to ensure alignment of SNP effect sizes. Within each individual nominated biomarker (exposure GWAS), We calculated Wald ratios for each extracted SNP by dividing the per-allele log-odds ratio (or beta) of that variant in the PD GWAS data by the log-odds ratio (or beta) of the same variant in the exposure data. For MR analysis, we applied inverse-variance weighted (IVW) and Egger methods to examine the relationship between the individual nominated biomarkers and PD. We studied the effects of pleiotropy and heterogeneity for each inference by undertaking sensitivity analyses. We performed reverse causality analyses to determine whether PD was causally linked to any of the nominated biomarkers. We only reported a relationship only when it was significant (raw p-value < 0.05) in both evaluations with IVW and Egger in the same direction and not having a reverse causation.

#### Discrimination ability of blood biomarkers

To evaluate the diagnostic accuracy of the nominated blood biomarkers, we trained and tested different machine learning algorithms to classify patients as PD cases or healthy controls at baseline. First, we employed lasso regression to eliminate superfluous variables that were not contributing to model accuracy. We then tested 4 different models on different combinations of biomarker and vital sign variables alongside the covariates that were kept constant across all models: age, sex, and BMI. We built the first model using α-synuclein, age, sex, and BMI. We built the second model using the blood biomarkers nominated by lasso (neutrophil %, monocyte, albumin, aspartate aminotransferase (AST), serum sodium) and the constant covariates. The third model used the same variables as the second, but three vital signs (systolic blood pressure in the standing position, heart rate in the standing position, and heart rate in the supine position) were added. The fourth model combined all the nominated variables and covariates used in the previous models. For each model, we tested 12 different binary classification algorithms using 5-fold cross validation (see supplemental materials for details). The best-performing algorithm was chosen according to the largest area under the curve (AUC), a measure of model performance.

#### The association between progression markers and MDS-UPDRS sub-scores

The associations between the MDS-UPDRS sub-scores (parts I, II and III) and the nominated progression markers were tested. We hypothesized that if the nominated progression markers were truly associated with progression, they may be associated with the MDS-UPDRS sub-scores at baseline among PD patients. Linear regression models were applied, adjusting for sex, age and BMI at the significance level of 0.05, two-tailed.

## Results

Table 1 shows the characteristics of the participants, the number of observations each year, the vital signs and the blood test results at baseline. In total, we had 603 participants and 3,592 data points in the current study. In the primary analysis, we nominated 10 differentiating biomarkers and 7 progression biomarkers among 19 vital sign associated outcomes and 30 different lab tests. Albumin and serum sodium were both differentiating and progression biomarkers. Table 2 presents the results for the 15 nominated biomarkers. (All the results can be found in Supplemental Table 1). Some of these biomarkers were correlated. When we analyzed the correlation coefficients of these measurements using the baseline data of controls, %WBC of lymphocyte and %WBC of neutrophil were highly correlated (correlation coefficient, rho = –0.95), indicating these two outcomes were effectively one (Supplemental Figure 1). There were also moderate correlations between supine and standing heart rates, cell counts and their %WBC for neutrophils and lymphocytes, and concentrations of serum sodium and chloride.

**Table 1:**
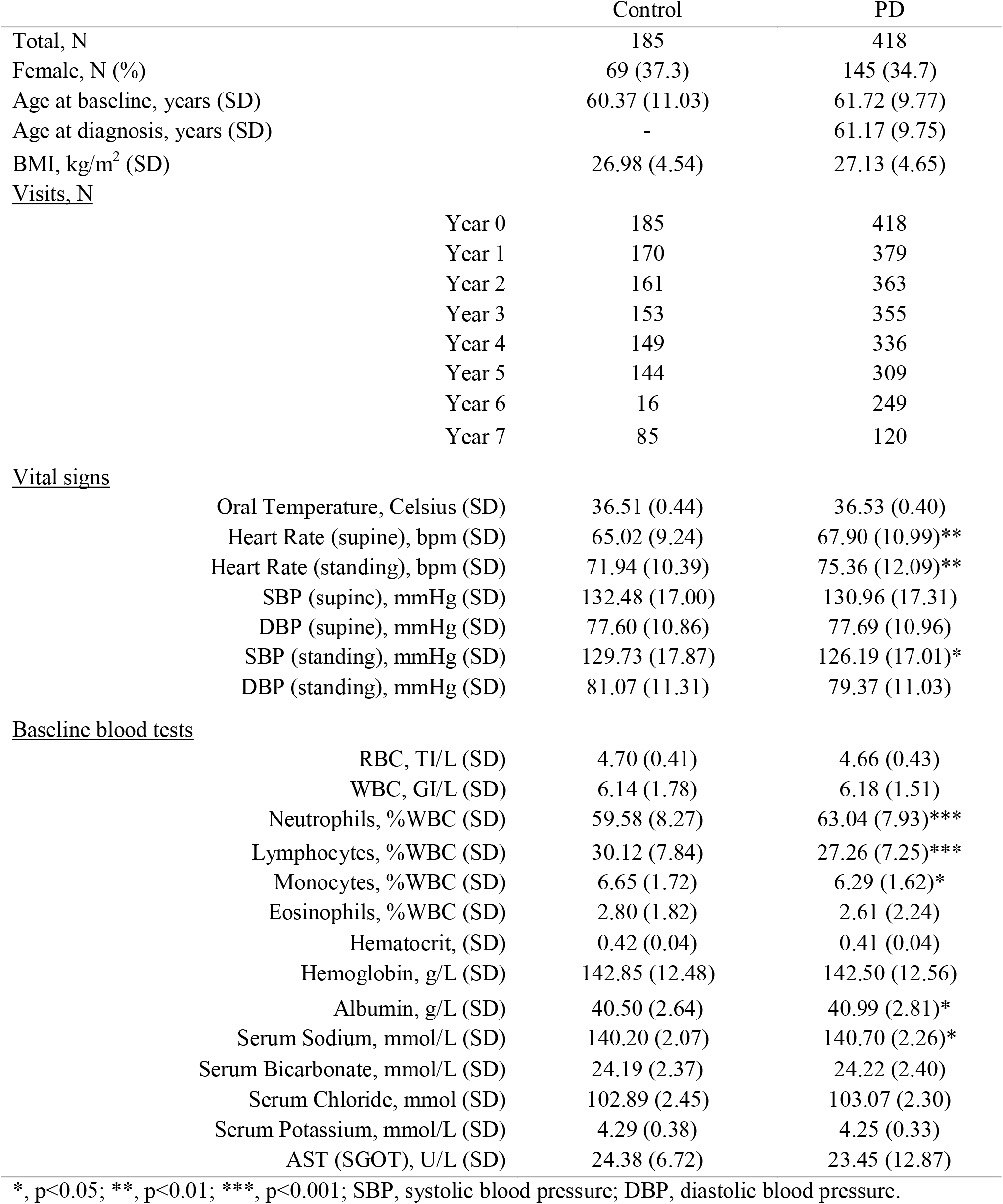
Baseline characteristics and selected vital signs and blood test results

**Table 2:**
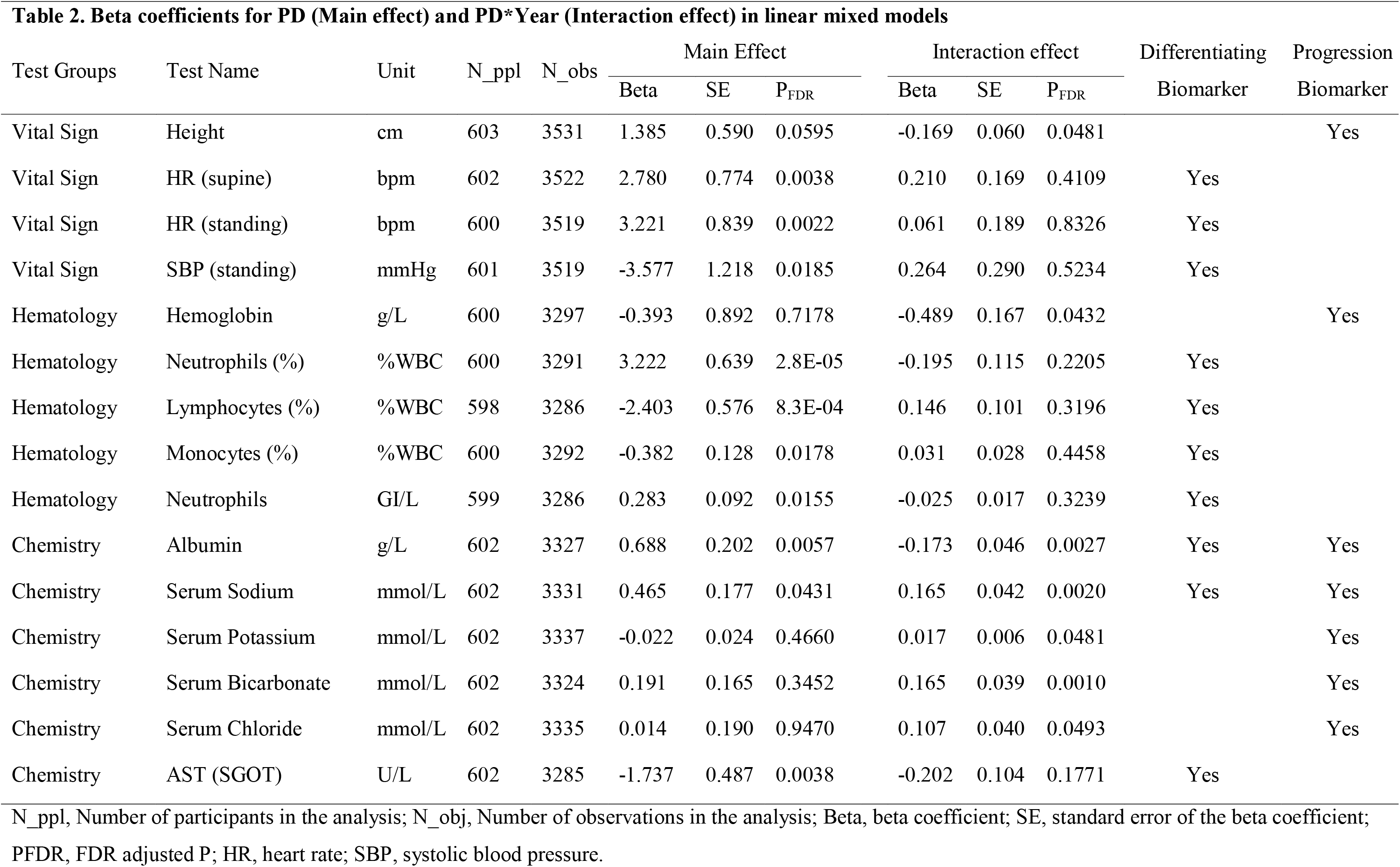
Beta coefficients for PD (Main effect) and PD*Year (Interaction effect) in linear mixed models

| Test Groups | Test Name | Unit | N_ppl | N_obs | Main Effect |  |  | Interaction effect |  |  | Differentiating Biomarker | Progression Biomarker |
| --- | --- | --- | --- | --- | --- | --- | --- | --- | --- | --- | --- | --- |
|  |  |  |  |  | Beta | SE | P<sub>FDR</sub> | Beta | SE | P<sub>FDR</sub> |  |  |
| Vital Sign | Height | cm | 603 | 3531 | 1.385 | 0.590 | 0.0595 | -0.169 | 0.060 | 0.0481 |  | Yes |
| Vital Sign | HR (supine) | bpm | 602 | 3522 | 2.780 | 0.774 | 0.0038 | 0.210 | 0.169 | 0.4109 | Yes |  |
| Vital Sign | HR (standing) | bpm | 600 | 3519 | 3.221 | 0.839 | 0.0022 | 0.061 | 0.189 | 0.8326 | Yes |  |
| Vital Sign | SBP (standing) | mmHg | 601 | 3519 | -3.577 | 1.218 | 0.0185 | 0.264 | 0.290 | 0.5234 | Yes |  |
| Hematology | Hemoglobin | g/L | 600 | 3297 | -0.393 | 0.892 | 0.7178 | -0.489 | 0.167 | 0.0432 |  | Yes |
| Hematology | Neutrophils (%) | % WBC | 600 | 3291 | 3.222 | 0.639 | 2.8E-05 | -0.195 | 0.115 | 0.2205 | Yes |  |
| Hematology | Lymphocytes (%) | % WBC | 598 | 3286 | -2.403 | 0.576 | 8.3E-04 | 0.146 | 0.101 | 0.3196 | Yes |  |
| Hematology | Monocytes (%) | % WBC | 600 | 3292 | -0.382 | 0.128 | 0.0178 | 0.031 | 0.028 | 0.4458 | Yes |  |
| Hematology | Neutrophils | GI/L | 599 | 3286 | 0.283 | 0.092 | 0.0155 | -0.025 | 0.017 | 0.3239 | Yes |  |
| Chemistry | Albumin | g/L | 602 | 3327 | 0.688 | 0.202 | 0.0057 | -0.173 | 0.046 | 0.0027 | Yes | Yes |
| Chemistry | Serum Sodium | mmol/L | 602 | 3331 | 0.465 | 0.177 | 0.0431 | 0.165 | 0.042 | 0.0020 | Yes | Yes |
| Chemistry | Serum Potassium | mmol/L | 602 | 3337 | -0.022 | 0.024 | 0.4660 | 0.017 | 0.006 | 0.0481 |  | Yes |
| Chemistry | Serum Bicarbonate | mmol/L | 602 | 3324 | 0.191 | 0.165 | 0.3452 | 0.165 | 0.039 | 0.0010 |  | Yes |
| Chemistry | Serum Chloride | mmol/L | 602 | 3335 | 0.014 | 0.190 | 0.9470 | 0.107 | 0.040 | 0.0493 |  | Yes |
| Chemistry | AST (SGOT) | U/L | 602 | 3285 | -1.737 | 0.487 | 0.0038 | -0.202 | 0.104 | 0.1771 | Yes |  |

N\_ppl, Number of participants in the analysis; N\_obj, Number of observations in the analysis; Beta, beta coefficient; SE, standard error of the beta coefficient; PFDR, FDR adjusted P; HR, heart rate; SBP, systolic blood pressure.

We observed a higher heart rate in the supine/standing position and lower systolic blood pressure in patients with PD. In the current study period, the differences between the supine and standing position in terms of blood pressure and heart rate were not significantly different, nor had they significantly increased over time, in patients with PD. These vital signs may suggest a slightly hypovolemic state among people with PD. A higher albumin concentration in the blood observed in PD cases aligned with this hypothesis, but we did not have adequate measurements to further confirm it. A negative change in height over time was another biomarker in the vital signs. Longitudinal changes in height in people with PD have rarely been reported, but given that slouching and scoliosis are common symptoms, they may explain the progressive lowering of height seen in the cases.

In the blood, the composition of white blood cells in PD cases was different to the composition of white blood cells in controls. The fraction of neutrophils had the strongest association with PD, and the association between PD and the fraction of lymphocytes may have been through the negative correlations between the fraction of lymphocytes and that of neutrophils. Among the chemistry tests, we nominated the concentration of AST as a differentiating biomarker, but we did not nominate alanine aminotransferase (ALT) as a differentiating biomarker. The discrepancy between the 2 enzymes may indicate the existence of reduced muscle volume in PD patients, even in the early phase. Further, we observed that the albumin and hemoglobin levels of PD patients were dropping faster than those of controls. This may suggest a nutrition deficit developing over time, but the average weight and its changing rate were not significantly different in PD patients during the study period. Finally, the composition of electrolytes changed over time among PD cases relative to controls. We adjusted for typical anti-PD medications, but we did not adjust for other drugs including laxative drugs, which might have influenced the results.

For the nominated biomarkers, we examined the mean difference between PD cases and controls at each visit after adjusting for sex, age and BMI (Figure 1); we also examined unadjusted boxplots of PD cases and controls at each visit (Supplemental Figure 2). These plots were useful to assess the chronological trends of the differences. All the nominated differentiating biomarkers were significantly different at baseline, suggesting the utility of these biomarkers for detecting PD in its early phase. Notably, the differences between PD cases and controls in terms of the differentiating biomarkers were relatively stable over time, indicating the robustness of these biomarkers against PD treatment and disease-associated changes. By contrast, the differences between PD cases and controls in terms of the nominated progression markers were changing over time, as we expected. We observed a sudden drop in the mean difference in albumin levels at the year-4 visit. We could not identify the particular reason for this phenomenon, but when we conducted the same analysis without using the year-4 visits, the model still suggested that the albumin level was both a differentiating biomarker and a progression biomarker.

**Figure 1:**
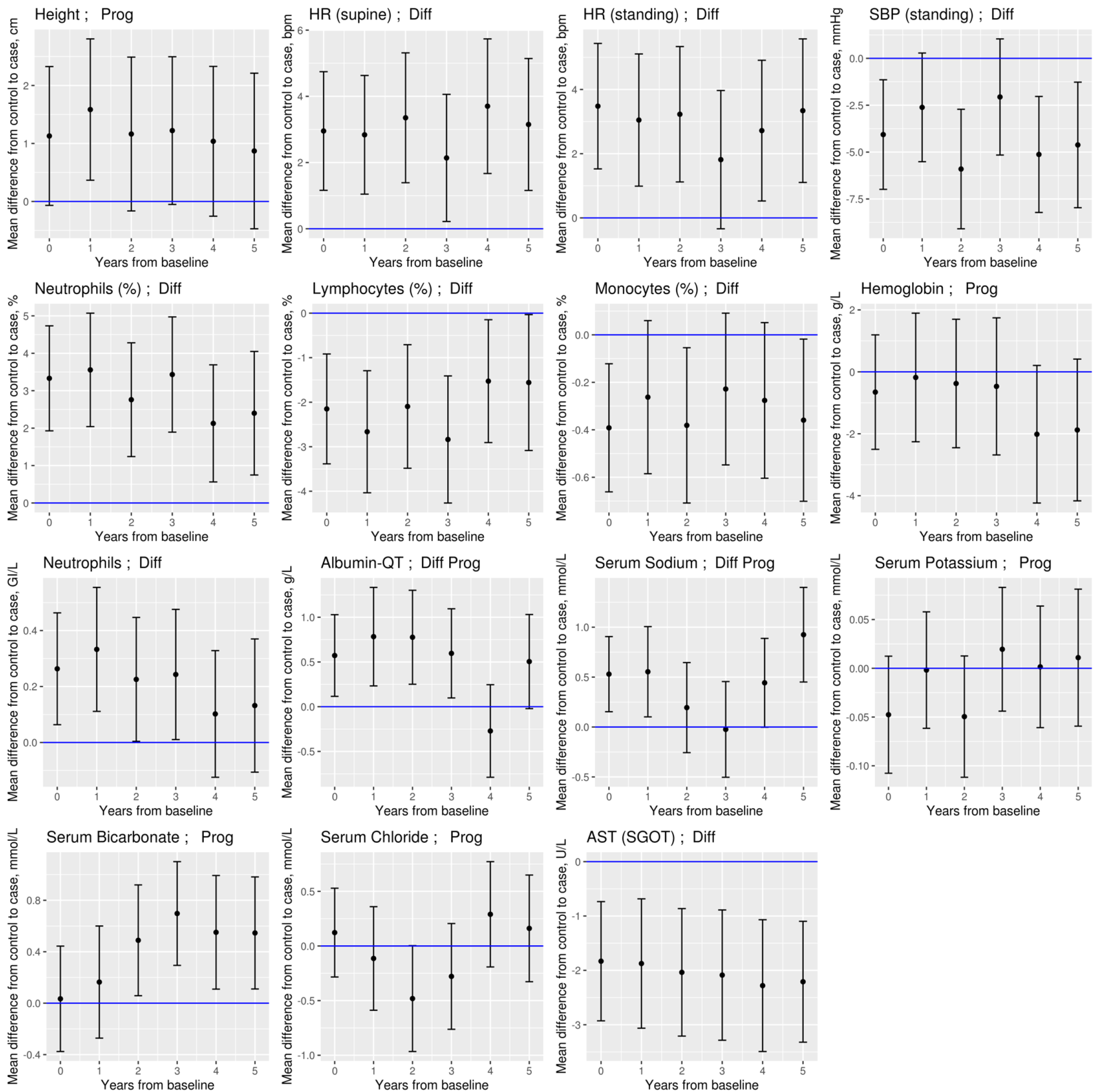
The mean differences between cases and controls at each visit. The Y-axis shows the mean differences between controls and cases. Bars indicate the 95% confidence interval. Age, sex, and BMI were adjusted. The blue horizontal line is 0. Prog = nominated as a progression biomarker; Diff = nominated as a differentiating biomarker.

**Figure 2:**
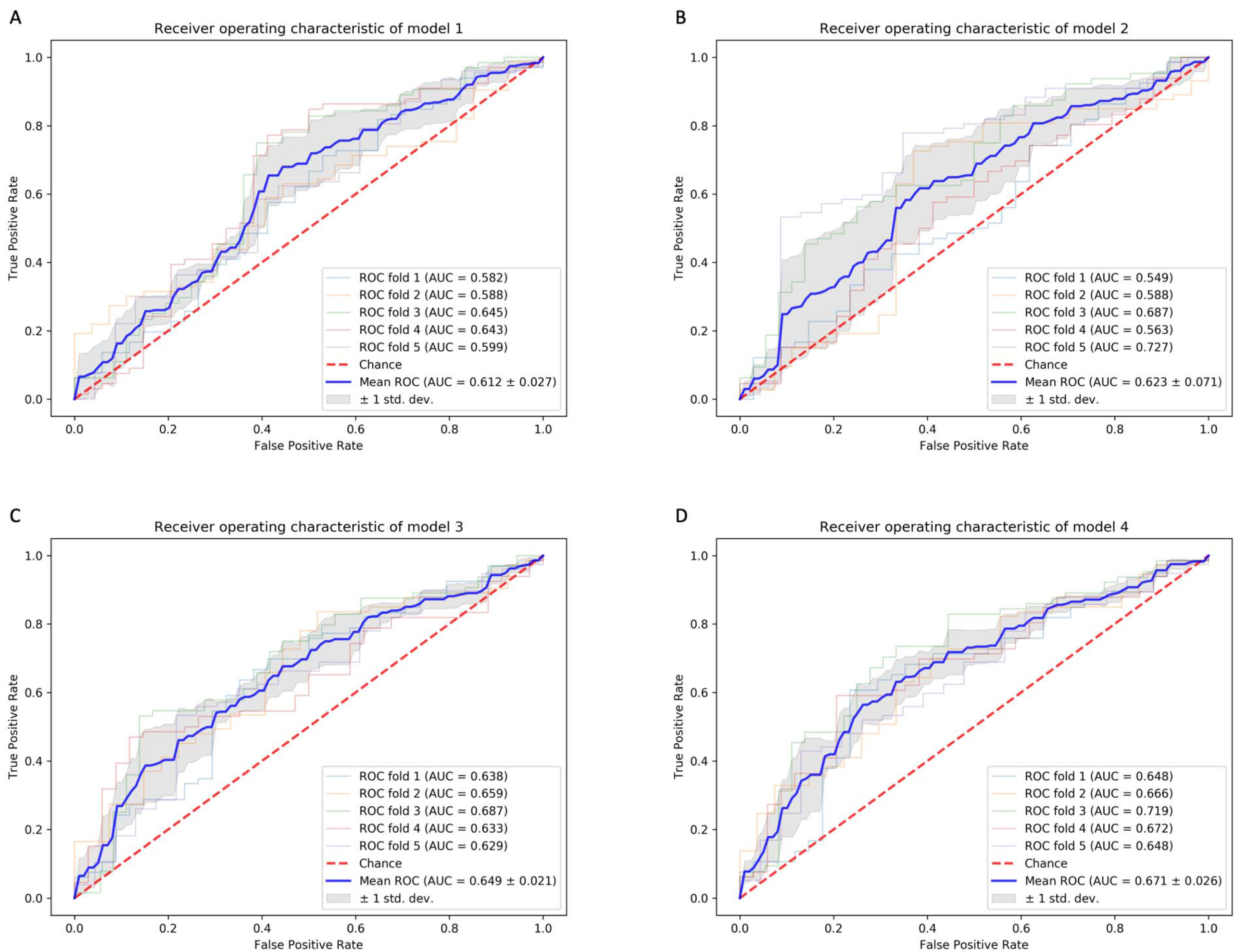
AUC of the prediction models. A. Model 1: Logistic regression. Variables: age(1),sex, BMI, and α-synuclein(2) B. Model 2: Adaptive Boosting classifier. Variables: age(1),sex, BMI(2), neutrophil %(3), monocyte %, albumin(5), AST, and serum sodium(4) C. Model 3: Linear Discriminant Analysis model. Variables: age, sex, BMI, neutrophil %(1), monocyte %(4), albumin(3), AST(5), serum sodium(2), systolic blood pressure in the standing position, heart rate in the standing position, and heart rate in the supine position(6) D. Model 4: Linear Discrimination Analysis model. Variables: age(1), sex, BMI(2), neutrophil %(3), monocyte %, albumin, AST(4), serum sodium(7), systolic blood pressure in the standing position(6), heart rate in the standing position, heart rate in the supine position, and α-synuclein(5) The numbers next to the variables indicate the rank of contribution to the model

The genetic approaches using LD score regression and Mendelian randomization did not indicate any association mechanism of shared genetic etiology between the nominated blood biomarkers and PD (Supplemental Table 3). Additionally, none of the assessed biomarkers showed any significant causal genetic relationship with PD risk or the presence of reverse causality (Supplemental Table 4). Sensitivity analyses did not find any sign of heterogeneity or horizontal pleiotropy.

When comparing the top performing predictive models, the accuracy of the model using basic variables (age, sex, BMI) plus α-synuclein (model 1) was similar to the model using the basic variables and blood variables (model 2) (Figure 2). The mean AUC increased when we added vital signs to model 2 (model 3) and further increased when we added α-synuclein (model 4). We performed feature ranking to determine the variables that contributed the most to each model. age, BMI, neutrophil (%), AST, albumin and serum sodium were nominated in multiple models.

Of the 7 progression biomarkers, 4 were associated with MDS-UPDRS sub-scores at baseline in PD. Sub-scores on MDS-UPDRS part I were significantly worse in patients with lower blood albumin levels, lower hemoglobin levels, or taller height. Sub-scores on MDS-UPDRS part III were worse in people with lower levels of serum bicarbonate (Supplemental Table 2). The other nominated progression biomarkers, however, were not associated with MDS-UPDRS sub-scores.

## Discussion

Among the biomarkers we screened, the neutrophil fraction in WBC had the strongest signal as a differentiating biomarker. In methylation data analyses, the estimated fraction of granulocytes has repeatedly been reported to be highly associated with PD status [13,14]. The current results suggest that this association may be driven by the fraction of neutrophils, which constitute the dominant portion of granulocytes. The neutrophil-lymphocyte ratio (NLR) has also been studied previously but its association with PD was inconclusive [15,16]. The NLR and neutrophil fraction in WBC were correlated in the current data, but the strength of the associations with PD was stronger for the neutrophil fraction (unadjusted *P* = 5.75E-07) than for the NLR (unadjusted *P* = 4.42E-06). Recently, a nested case-control study screening peripheral immune biomarkers in a large population cohort in Sweden reported that a lower concentration of white blood cells, especially lymphocytes was associated with an increased incidence of PD [17]. In the current study, the direction of the association between lymphocyte counts and PD was concordant with this finding but did not reach the FDR threshold (*P* = 0.053). Moreover, the lymphocyte count in the current data was moderately associated with the strongest predictor, neutrophil fraction of WBC (rho = –0.57). Further investigations are needed to identify the most important measurement representing the differences between PD cases and controls in terms of hematologic profiles. The reason for the change in white blood cell composition is not well known, although inflammation is hypothesized to be a major pathogenic factor for PD [18]. Inflammatory cytokines may play a role. A nested case-control study of a prospective cohort reported that higher concentrations of Interleukin-6 were found in the blood collected on average 4.3 years before the diagnosis [19,20]. It is also interesting that a change in the gut bacterial population has been reported in patients with PD as well as those with REM sleep behavior disorders [20], although studies of the association between the hematology profile and the gut bacterial population are rare. We conducted an ad-hoc test to assess the association between the prevalence of constipation and the neutrophil fraction in WBC in PD patients at baseline, but the association was null (*P* = 0.92).

Based on the analyses applying genetic approaches which did not suggest any shared genetic etiology or causality of the association mechanisms, we do not have any evidence that the associations in the current study were more than the disease associated changes. Still, studying the reasons behind these associations is warranted because it may lead to the identification of important disease mechanisms or the better management of patients.

The prediction models using basic variables and biomarkers had relatively low AUCs and they were not useful for differentiating PD patients from healthy controls in daily practice. Nevertheless, the ranking of variables contributing to the prediction models may be useful to prioritize the biomarkers for further research.

The current study has 3 strengths. First, we leveraged the longitudinal data not only to obtain higher power in detecting differentiating biomarkers but also as an opportunity to find progression biomarkers. Second, the vital signs and laboratory tests we screened were commonly obtained in a routine clinical setting. Although this study was exploratory, we demonstrated the potential value of these measurements to assess the differences between PD cases and controls and to assess the disease progression. Although these biomarkers were substantially overlapped between cases and controls and may not be useful for diagnosis, the study provided some hypotheses explaining the differences which can be confirmed in everyday practice. Third, unlike many other biomarker studies, in which PD patients at various stages were included and compared with controls, this study included PD patients who were all at an early disease stage and untreated at baseline. In this setting, we were able to untangle the impact of treatment and PD-associated changes in the progression stage. It is important to note that all the differentiating biomarkers were confirmed to be significantly different in PD cases than in controls at baseline.

The major limitation is that we did not approach the individual mechanisms underlying the differences between PD cases and controls in terms of these biomarkers. As similar studies of this size and span are rare, the results from this effort will be useful to generate hypotheses that ought to be tested further. Another limitation relates to the setting of the study: Most of the cases in the study were in relatively early stages of the disease. Longer observations will be necessary to identify progression markers relevant to the later stage of the disease.

In conclusion, we screened vital signs and common blood test measurements over more than 5 years to identify differentiating/progression biomarkers of PD. A total of 16 measurements were nominated. These associations should be studied further.

## Data Availability

The participants in this study included 418 patients with PD (cases) and 185 individuals without any neurological disease (controls) from the PPMI. Detailed information regarding the PPMI is available at www.ppmi-info.org.

## Acknowledgments

This study was supported by the Intramural Research Program the National Institute on Aging (Z01 AG000949 02), and the Michael J. Fox Foundation for Parkinson’s Research. This research was supported in part by the Intramural Research Program of the National Institutes of Health, National Institute of Neurological Disorders and Stroke (project number: 1ZIANS003154).

Data used in preparation of this article were obtained from the MJFF sponsored PPMI database (www.ppmi-info.org/data). For up to date information on these studies, visit www.ppmi-info.org. PPMI – a public-private partnership – is funded by The Michael J. Fox Foundation for Parkinson’s Research and funding partners, including AbbVie, Allergan, Avid Radiopharmaceuticals, Biogen, BioLegend, Bristol-Myers Squibb, Celgene, Denali Incorporated, GE Healthcare, Genentech, GlaxoSmithKline, Eli Lilly and Company, Lundbeck, Merck & Co., Meso Scale Discovery, Pfizer, Piramal, Prevail Therapeutics, Roche, Sanofi Genzyme, Servier Laboratories, Takeda, Teva, UCB, Verily, Voyager Therapeutics, and Golub Capital (www.ppmi-info.org/fundingpartners).

## Conflict of Interest

The authors have no conflict of interest to report.

## Supplemental material

**Supplemental Figure 1:**
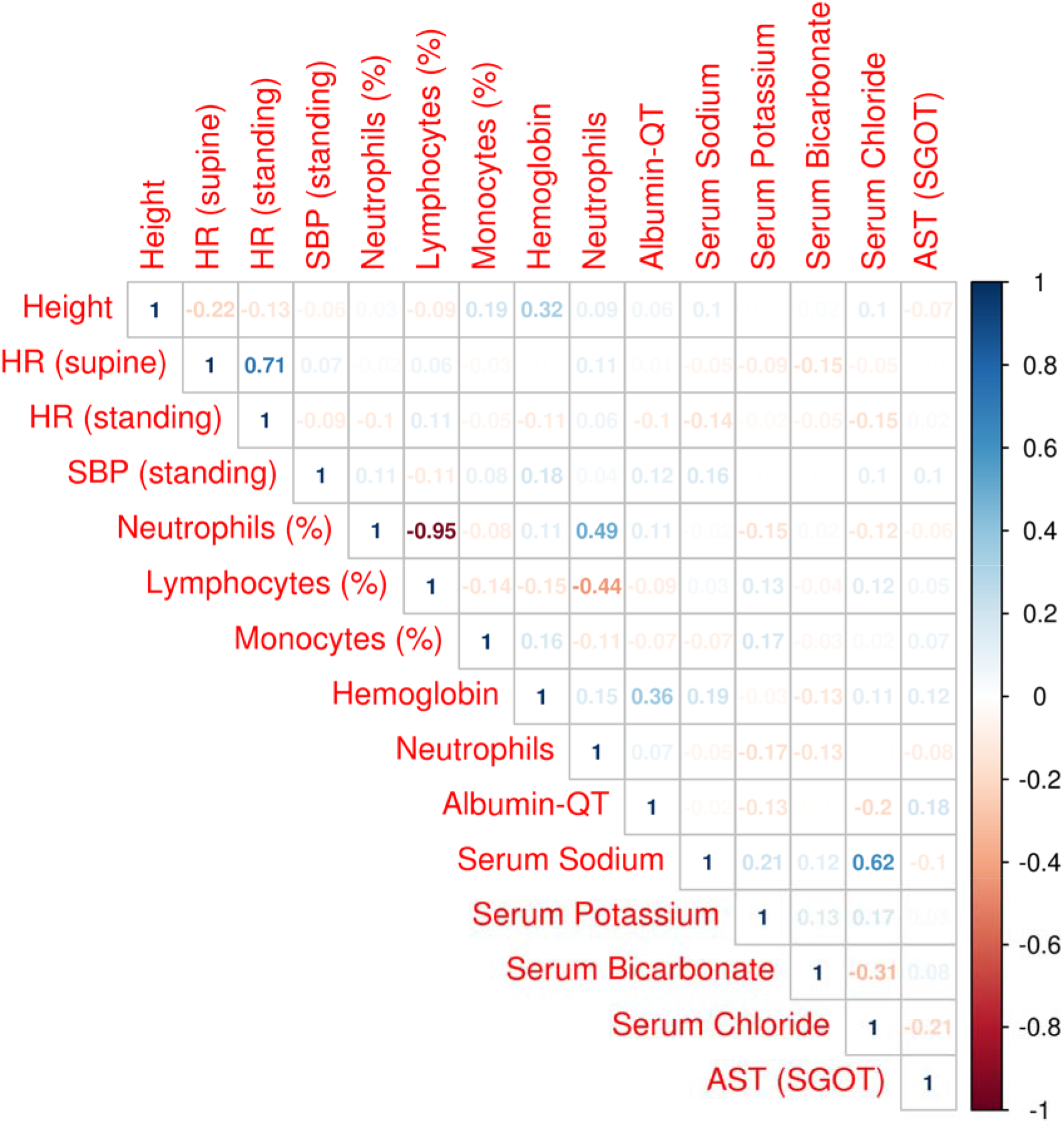
Correlation coefficients of the nominated biomarkers in the controls at baseline. Some of the nominated biomarkers were highly correlated.

**Supplemental Figure 2:**
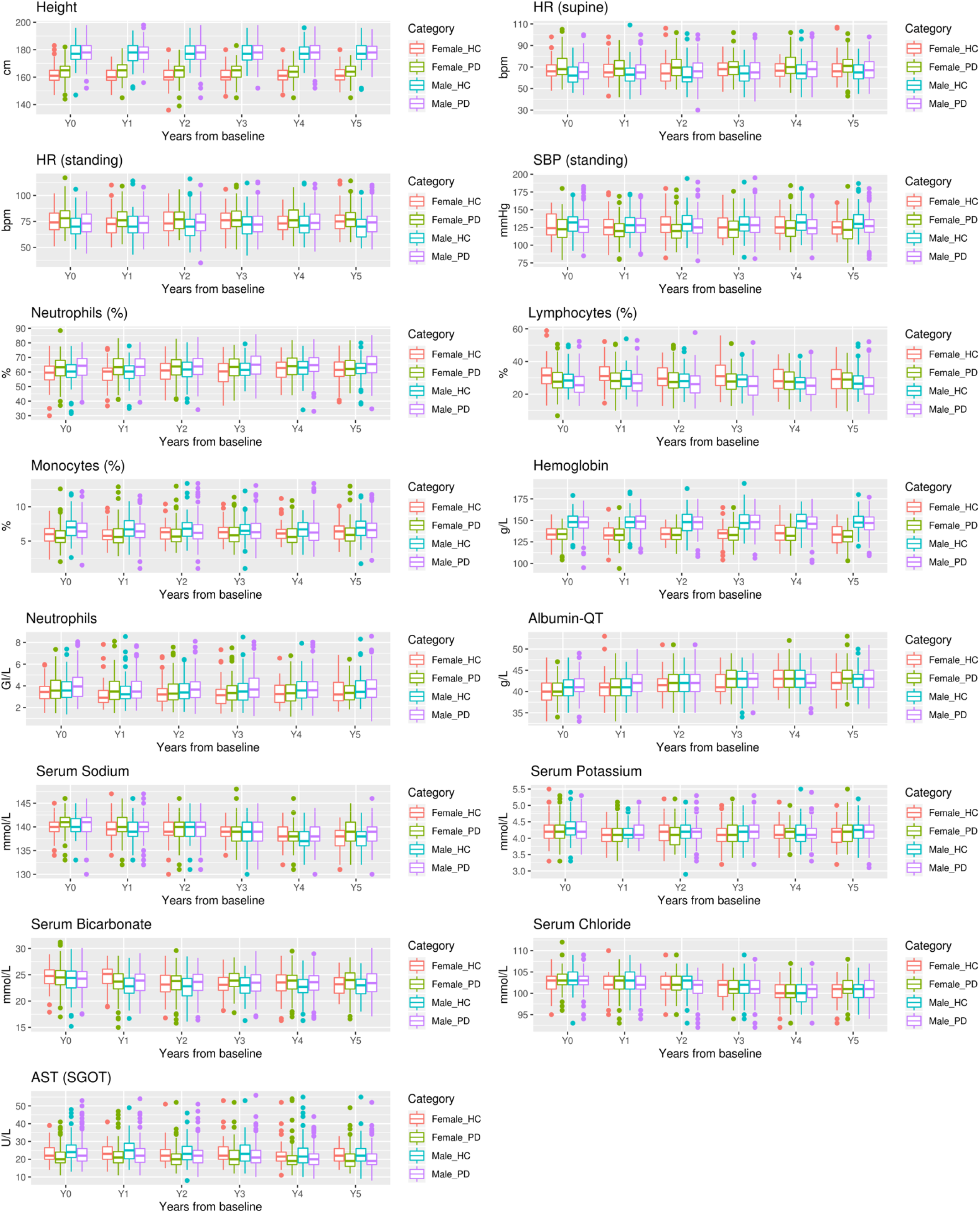
Boxplots of the unadjusted values of the biomarkers at each visit, sex stratified.

**Supplemental Table 1:** All results of the primary analysis

| Test Groups | Test Name | Unit | N_ppl | N_obs | Main Effect |  |  |  | Interaction Effect |  |  |  |
| --- | --- | --- | --- | --- | --- | --- | --- | --- | --- | --- | --- | --- |
|  |  |  |  |  | Beta | SE | P<sub>raw</sub> | P<sub>FDR</sub> | Beta | SE | P<sub>raw</sub> | P<sub>FDR</sub> |
| VITAL | Weight | kg | 603 | 3527 | 1.34 | 1.25 | 0.284 | 0.376 | -0.30 | 0.13 | 0.020 | 0.107 |
| VITAL | Height | cm | 603 | 3531 | 1.39 | 0.59 | 0.019 | 0.060 | -0.17 | 0.06 | 0.005 | 0.048 |
| VITAL | Temperature | Celsius | 601 | 3509 | 0.02 | 0.03 | 0.599 | 0.667 | 0.00 | 0.01 | 0.579 | 0.692 |
| VITAL | HR (supine) | bpm | 602 | 3522 | 2.78 | 0.77 | 3.5E-04 | 0.004 | 0.21 | 0.17 | 0.215 | 0.411 |
| VITAL | SBP (supine) | mmHg | 602 | 3522 | -1.60 | 1.19 | 0.180 | 0.276 | 0.54 | 0.29 | 0.060 | 0.177 |
| VITAL | DBP (supine) | mmHg | 602 | 3523 | 0.07 | 0.77 | 0.923 | 0.947 | 0.14 | 0.16 | 0.376 | 0.526 |
| VITAL | HR (standing) | bpm | 600 | 3519 | 3.22 | 0.84 | 1.3E-04 | 0.002 | 0.06 | 0.19 | 0.745 | 0.833 |
| VITAL | SBP (standing) | mmHg | 601 | 3519 | -3.58 | 1.22 | 0.003 | 0.019 | 0.26 | 0.29 | 0.363 | 0.523 |
| VITAL | DBP (standing) | mmHg | 601 | 3522 | -1.22 | 0.79 | 0.122 | 0.199 | 0.05 | 0.18 | 0.784 | 0.833 |
| VITAL | MBP (supine) | mmHg | 602 | 3522 | -0.56 | 0.83 | 0.496 | 0.565 | 0.29 | 0.18 | 0.111 | 0.258 |
| VITAL | MBP (standing) | mmHg | 601 | 3522 | -2.05 | 0.85 | 0.016 | 0.060 | 0.12 | 0.19 | 0.525 | 0.644 |
| VITAL | SBP (difference) | mmHg | 600 | 3507 | -1.90 | 0.86 | 0.027 | 0.077 | -0.25 | 0.22 | 0.240 | 0.436 |
| VITAL | DBP (difference) | mmHg | 600 | 3511 | -1.05 | 0.54 | 0.054 | 0.114 | -0.10 | 0.13 | 0.442 | 0.586 |
| VITAL | HR (difference) | mmHg | 599 | 3510 | 0.63 | 0.49 | 0.201 | 0.297 | -0.24 | 0.12 | 0.045 | 0.177 |
| VITAL | SBP (ratio) |  | 600 | 3514 | -0.01 | 0.01 | 0.044 | 0.097 | 0.00 | 0.00 | 0.282 | 0.446 |
| VITAL | DBP (ratio) |  | 601 | 3508 | -0.01 | 0.01 | 0.074 | 0.135 | 0.00 | 0.00 | 0.360 | 0.523 |
| VITAL | HR (ratio) |  | 600 | 3503 | 0.01 | 0.01 | 0.490 | 0.565 | 0.00 | 0.00 | 0.065 | 0.177 |
| VITAL | MBP (difference) | mmHg | 600 | 3512 | -1.39 | 0.58 | 0.016 | 0.060 | -0.14 | 0.14 | 0.310 | 0.475 |
| VITAL | MBP (ratio) |  | 600 | 3513 | -0.01 | 0.01 | 0.041 | 0.097 | 0.00 | 0.00 | 0.274 | 0.446 |
| HEMAT | Monocytes | GI/L | 599 | 3285 | -0.02 | 0.01 | 0.066 | 0.130 | 0.00 | 0.00 | 0.725 | 0.833 |
| HEMAT | Eosinophils | GI/L | 599 | 3270 | -0.01 | 0.01 | 0.092 | 0.155 | 0.00 | 0.00 | 0.779 | 0.833 |
| HEMAT | Basophils | GI/L | 600 | 3276 | 0.00 | 0.00 | 0.892 | 0.947 | 0.00 | 0.00 | 0.883 | 0.901 |
| HEMAT | Platelets | GI/L | 600 | 3267 | -7.75 | 4.57 | 0.090 | 0.155 | 1.27 | 0.66 | 0.054 | 0.177 |
| HEMAT | Neutrophils (%) | % | 600 | 3291 | 3.22 | 0.64 | 5.8E-07 | 2.8E-05 | -0.20 | 0.11 | 0.089 | 0.220 |
| HEMAT | Lymphocytes (%) | % | 598 | 3286 | -2.40 | 0.58 | 3.4E-05 | 0.001 | 0.15 | 0.10 | 0.150 | 0.320 |
| HEMAT | Monocytes (%) | % | 600 | 3292 | -0.38 | 0.13 | 0.003 | 0.018 | 0.03 | 0.03 | 0.273 | 0.446 |
| HEMAT | Eosinophils (%) | % | 600 | 3278 | -0.27 | 0.11 | 0.019 | 0.060 | 0.01 | 0.02 | 0.778 | 0.833 |
| HEMAT | Basophils (%) | % | 599 | 3276 | -0.04 | 0.03 | 0.147 | 0.232 | 0.01 | 0.01 | 0.090 | 0.220 |
| HEMAT | Hematocrit |  | 600 | 3294 | 0.00 | 0.00 | 0.267 | 0.363 | 0.00 | 0.00 | 0.065 | 0.177 |
| HEMAT | RBC | TI/L | 600 | 3299 | -0.03 | 0.03 | 0.338 | 0.436 | -0.01 | 0.01 | 0.056 | 0.177 |
| HEMAT | Hemoglobin | g/L | 600 | 3297 | -0.39 | 0.89 | 0.659 | 0.718 | -0.49 | 0.17 | 0.004 | 0.043 |
| HEMAT | WBC | GI/L | 598 | 3284 | 0.10 | 0.12 | 0.412 | 0.505 | -0.02 | 0.02 | 0.278 | 0.446 |
| HEMAT | Neutrophils | GI/L | 599 | 3286 | 0.28 | 0.09 | 0.002 | 0.015 | -0.02 | 0.02 | 0.159 | 0.324 |
| HEMAT | Lymphocytes | GI/L | 598 | 3287 | -0.11 | 0.04 | 0.012 | 0.053 | 0.00 | 0.01 | 0.515 | 0.644 |
| CHEMI | Total Bilirubin | umol/L | 602 | 3275 | 0.81 | 0.39 | 0.037 | 0.096 | -0.05 | 0.06 | 0.396 | 0.538 |
| CHEMI | Serum Glucose | mmol/L | 601 | 3298 | 0.12 | 0.06 | 0.041 | 0.097 | -0.03 | 0.01 | 0.031 | 0.140 |
| CHEMI | Total Protein | g/L | 602 | 3340 | 0.71 | 0.30 | 0.019 | 0.060 | -0.10 | 0.07 | 0.119 | 0.264 |
| CHEMI | Albumin-QT | g/L | 602 | 3327 | 0.69 | 0.20 | 0.001 | 0.006 | -0.17 | 0.05 | 0.000 | 0.003 |
| CHEMI | Alkaline Phosphatase-QT | U/L | 602 | 3327 | -0.10 | 1.44 | 0.947 | 0.947 | 0.53 | 0.24 | 0.029 | 0.140 |
| CHEMI | Serum Sodium | mmol/L | 602 | 3331 | 0.47 | 0.18 | 0.009 | 0.043 | 0.17 | 0.04 | 0.000 | 0.002 |
| CHEMI | Serum Potassium | mmol/L | 602 | 3337 | -0.02 | 0.02 | 0.371 | 0.466 | 0.02 | 0.01 | 0.006 | 0.048 |
| CHEMI | Serum Bicarbonate | mmol/L | 602 | 3324 | 0.19 | 0.16 | 0.247 | 0.345 | 0.17 | 0.04 | 0.000 | 0.001 |
| CHEMI | Serum Chloride | mmol/L | 602 | 3335 | 0.01 | 0.19 | 0.942 | 0.947 | 0.11 | 0.04 | 0.007 | 0.049 |
| CHEMI | Calcium (EDTA) | mmol/L | 602 | 3332 | 0.02 | 0.01 | 0.028 | 0.077 | 0.00 | 0.00 | 0.984 | 0.984 |
| CHEMI | Creatinine (Rate Blanked) | umol/L | 601 | 3337 | 0.86 | 1.15 | 0.455 | 0.544 | 0.05 | 0.20 | 0.799 | 0.833 |
| CHEMI | ALT (SGPT) | U/L | 602 | 3294 | -1.35 | 0.74 | 0.070 | 0.131 | -0.13 | 0.17 | 0.460 | 0.593 |
| CHEMI | AST (SGOT) | U/L | 602 | 3285 | -1.74 | 0.49 | 3.8E-04 | 0.004 | -0.20 | 0.10 | 0.052 | 0.177 |
| CHEMI | Urea Nitrogen | mmol/L | 601 | 3326 | 0.15 | 0.12 | 0.206 | 0.297 | 0.03 | 0.02 | 0.218 | 0.411 |
| CHEMI | Serum Uric Acid | umol/L | 602 | 3342 | -9.59 | 5.08 | 0.060 | 0.121 | -2.43 | 0.92 | 0.009 | 0.054 |

N\_ppl, Number of participants in the analysis; N\_obj, Number of observations in the analysis; Beta, beta coefficient; SE, standard error of the beta coefficient; P<sub>FDR</sub>, FDR adjusted P; HR, heart rate; SBP, systolic blood pressure; MBP, mean blood pressure; HEMAT, Hematology test; CHEMI, chemistry test.

**Supplemental Table 2:** Associations between the progression biomarkers and the MDS-UPDRS sub-scores of patients with PD at baseline

| Test Group | Test Name | Unit | Outcome | Beta | SE | Praw |
| --- | --- | --- | --- | --- | --- | --- |
| Vital Sign | Height | cm | UPDRS1 | 0.066 | 0.029 | 0.023 |
| Vital Sign | Height | cm | UPDRS2 | 0.054 | 0.030 | 0.075 |
| Vital Sign | Height | cm | UPDRS3 | 0.092 | 0.063 | 0.145 |
| Hematology | Hemoglobin | g/L | UPDRS1 | -0.064 | 0.019 | 0.001 |
| Hematology | Hemoglobin | g/L | UPDRS3 | -0.051 | 0.043 | 0.231 |
| Hematology | Hemoglobin | g/L | UPDRS2 | -0.005 | 0.020 | 0.805 |
| Chemistry | Albumin-QT | g/L | UPDRS1 | -0.183 | 0.076 | 0.016 |
| Chemistry | Albumin-QT | g/L | UPDRS2 | -0.078 | 0.079 | 0.327 |
| Chemistry | Albumin-QT | g/L | UPDRS3 | -0.011 | 0.166 | 0.946 |
| Chemistry | Serum Sodium | mmol/L | UPDRS3 | -0.266 | 0.195 | 0.173 |
| Chemistry | Serum Sodium | mmol/L | UPDRS2 | -0.099 | 0.093 | 0.288 |
| Chemistry | Serum Sodium | mmol/L | UPDRS1 | -0.091 | 0.090 | 0.311 |
| Chemistry | Serum Potassium | mmol/L | UPDRS1 | 0.140 | 0.604 | 0.817 |
| Chemistry | Serum Potassium | mmol/L | UPDRS2 | 0.055 | 0.627 | 0.930 |
| Chemistry | Serum Potassium | mmol/L | UPDRS3 | -0.007 | 1.316 | 0.996 |
| Chemistry | Serum Bicarbonate | mmol/L | UPDRS3 | -0.406 | 0.186 | 0.029 |
| Chemistry | Serum Bicarbonate | mmol/L | UPDRS2 | -0.100 | 0.089 | 0.261 |
| Chemistry | Serum Bicarbonate | mmol/L | UPDRS1 | -0.057 | 0.086 | 0.510 |
| Chemistry | Serum Chloride | mmol/L | UPDRS1 | 0.030 | 0.087 | 0.726 |
| Chemistry | Serum Chloride | mmol/L | UPDRS2 | 0.009 | 0.090 | 0.917 |
| Chemistry | Serum Chloride | mmol/L | UPDRS3 | 0.015 | 0.189 | 0.935 |

Highlight, FDR adjusted $P < 0.05$ . "

**Supplemental Table 3:** Genetic correlations between Parkinson’s disease and the nominated biomarkers

| Phenotype 1 | Phenotype 2 | rg | SE | P | h2_obs | h2_obs<br>_SE | h2_int | h2_int_SE | gcov_<br>int | gcov_<br>int_SE |
| --- | --- | --- | --- | --- | --- | --- | --- | --- | --- | --- |
| Parkinson | Hemoglobin concentration | 0.036 | 0.032 | 0.265 | 0.158 | 0.015 | 1.098 | 0.023 | -0.009 | 0.006 |
| Parkinson | Lymphocyte counts | -0.027 | 0.030 | 0.365 | 0.211 | 0.017 | 1.060 | 0.017 | 0.007 | 0.006 |
| Parkinson | Lymphocyte percentage | -0.016 | 0.033 | 0.616 | 0.161 | 0.013 | 1.048 | 0.014 | 0.002 | 0.006 |
| Parkinson | Monocyte percentage | -0.040 | 0.032 | 0.215 | 0.185 | 0.023 | 1.102 | 0.030 | 0.002 | 0.007 |
| Parkinson | Neutrophil count | 0.002 | 0.032 | 0.951 | 0.177 | 0.021 | 1.048 | 0.018 | 0.003 | 0.006 |
| Parkinson | Neutrophil percentage | 0.032 | 0.034 | 0.338 | 0.155 | 0.013 | 1.038 | 0.015 | -0.001 | 0.006 |

rg, genetic correlation; SE, standard error; h<sup>2</sup> obs, narrow-sense heritability observed; h<sup>2</sup> int, narrow-sense heritability intercept, gcov\_int, cross-trait intercept.

**Supplemental Table 4:** Two-sample Mendelian randomization between Parkinson’s disease and the nominated biomarkers

| Exposure | Outcome | no. of SNPs | IVW |  |  | MR Egger |  |  |
| --- | --- | --- | --- | --- | --- | --- | --- | --- |
|  |  |  | beta | se | P | beta | se | P |
| Hemoglobin concentration | Parkinson | 110 | -0.03 | 0.08 | 0.75 | -0.02 | 0.16 | 0.91 |
| Parkinson | Hemoglobin concentration | 62 | -0.01 | 0.01 | 0.14 | -0.03 | 0.02 | 0.10 |
| Lymphocyte counts | Parkinson | 141 | -0.11 | 0.06 | 0.06 | 0.25 | 0.15 | 0.11 |
| Parkinson | Lymphocyte counts | 62 | -0.01 | 0.01 | 0.23 | 0.00 | 0.02 | 0.82 |
| Lymphocyte percentage | Parkinson | 116 | -0.04 | 0.10 | 0.68 | 0.12 | 0.28 | 0.67 |
| Parkinson | Lymphocyte percentage | 62 | 0.00 | 0.01 | 0.82 | 0.06 | 0.02 | 0.02 |
| Monocyte percentage | Parkinson | 161 | -0.01 | 0.05 | 0.76 | -0.02 | 0.09 | 0.86 |
| Parkinson | Monocyte percentage | 62 | -0.01 | 0.01 | 0.57 | -0.01 | 0.02 | 0.51 |
| Neutrophil count | Parkinson | 120 | -0.01 | 0.07 | 0.90 | -0.19 | 0.15 | 0.21 |
| Parkinson | Neutrophil count | 62 | -0.01 | 0.01 | 0.33 | -0.07 | 0.03 | 0.02 |
| Neutrophil percentage | Parkinson | 116 | 0.06 | 0.08 | 0.49 | 0.14 | 0.20 | 0.49 |
| Parkinson | Neutrophil percentage | 62 | 0.00 | 0.01 | 0.82 | -0.06 | 0.02 | 0.02 |

## Model specifications

### Linear mixed model

#### For height, weight, and BMI

OUTCOME ∽ PD*START + FEMALE + AGEatBL + AMTDN + LDOPA + MAOBI + PRMXL + RPNRL +(START|PATNO)

#### Outcome other than above

OUTCOME ∽ PD*START + FEMALE + AGEatBL + BMI + AMTDN + LDOPA + MAOBI + PRMXL + RPNRL +(START|PATNO)

#### Linear model for each visit

OUTCOME ∽ PD + FEMALE + AGEatBL + BMI

✶ PD, indicator variable for PD; AGEatBL, age at baseline; AMTDN, amantadine dose; MAOBI, MAO-B inhibitor dose;LDOPA, Levodopa dose; PRMXL, pramipexole dose; RPNRL, ropinirol dose.

## Prediction models and its selection

| Function Name | Package (python) |
| --- | --- |
| LogisticRegression | sklearn.linear_model |
| SGDClassifier | sklearn.linear_model |
| RandomForestClassifier | sklearn.ensemble |
| AdaBoostClassifier | sklearn.ensemble |
| GradientBoostingClassifier | sklearn.ensemble |
| BaggingClassifier | sklearn.ensemble |
| SVC | sklearn.svm |
| MLPClassifier | sklearn.neural_network |
| KNeighborsClassifier | sklearn.neighbors |
| LinearDiscriminantAnalysis | sklearn.discriminant_analysis |
| QuadraticDiscriminantAnalysis | sklearn.discriminant_analysis |
| XGBClassifier | xgboost |

The prediction abilities of the differentiating biomarkers were evaluated using baseline data. All algorithms were tested with 5-fold cross validation and one with the largest mean area under the curve were selected for reporting.

Model 1 variables: AGEatBL, FEMALE, BMI, α-synuclein

Model 2 variables: AGEatBL, FEMALE, BMI, Neutrophil %, Monocyte %, Albumin, AST, Serum Sodium

Model 3 variables: AGEatBL, FEMALE, BMI, Neutrophil %, Monocyte %, Albumin, AST, Serum Sodium, systolic blood pressure at standing, heart rate at standing position, and heart rate at supine position

Model 4 variables: AGEatBL, FEMALE, BMI, Neutrophil %, Monocyte %, Albumin, AST, Serum Sodium, systolic blood pressure at standing, heart rate at standing position, and heart rate at supine position, aSYN.α-synuclein

* AGEatBL, age at baseline

## Notes

### Competing Interest Statement

The authors have declared no competing interest.

### Clinical Trial

NCT01141023

### Funding Statement

Hirotaka Iwaki: Grants, Michael J Fox Foundation, Mike A. Nalls: Consultancies, Lysosomal Therapies Inc., Vivid Genomics Inc., Kleiner Perkins Caufield & Byers, Neuron23, Inc., Michael J. Fox Foundation

